# Characterizing the performance of a DIY air filter

**DOI:** 10.1101/2022.01.09.22268972

**Authors:** Rachael Dal Porto, Monet N. Kunz, Theresa Pistochini, Richard L. Corsi, Christopher D. Cappa

## Abstract

Air filtration serves to reduce concentrations of particles in indoor environments. Most standalone, also referred to as portable or in-room, air filtration systems use HEPA filters, and cost generally scales with the clean air delivery rate. A “do-it-yourself” lower-cost alternative, known as the Corsi-Rosenthal Box, that uses MERV-13 filters coupled with a box fan has been recently proposed, but lacks systematic performance characterization. We have characterized the performance of a five-panel Corsi-Rosenthal air filter. Measurements of size-resolved and overall decay rates of aerosol particles larger than 0.5 microns emitted into rooms of varying size with and without the air filter allowed for determination of the apparent clean air delivery rate—both as a function of size and integrated across particle sizes. The measurements made in the different rooms produced similar results, demonstrating the robustness of the method used. The size-integrated apparent clean air delivery rate increases with fan speed, from about 600 to 850 ft^3^ min^−1^ (1019 to 1444 m^3^ h^−1^). Overall, our results demonstrate that the Corsi-Rosenthal filter efficiently reduces suspended particle concentrations in indoor environments.

**One Sentence Summary:** A DIY air cleaner can effectively reduce aerosols in indoor spaces.

## 1 Introduction

Filtration is a robust and widely used method to reduce particle concentrations in indoor environments (Curtius, Granzin and Schrod 2021; Kelly and Fussell 2019; McNamara et al. 2017; MillerLeiden et al. 1996). Particle filters can be embedded in ventilation systems or added as stand-alone, portable units within rooms (Alavy and Siegel 2020; Shaughnessy and Sextro 2006). Filters vary widely in their efficiency and are characterized by the minimum efficiency reporting value (MERV), with the highest efficiency filters referred to as high efficiency particulate air (HEPA) filters (ASHRAE 2017). Filter efficiency varies with particle size, and HEPA filters remove at least 99.97% of particles having diameters of 0.3 microns, which is typically where the minimum filter efficiency occurs. While ventilation systems rarely use HEPA filters, owing to the accompanying large pressure drop and space requirements, most commercial in-room filtration systems rely on HEPA filters (Shaughnessy and Sextro 2006). Various studies support the benefits of portable HEPA filtration for reducing aerosol concentrations from many sources, including reducing risks of COVID-19 transmission. For example, Liu et al. (2021) reviewed portable HEPA air cleaners and concluded that such air cleaners have “potential to eliminate airborne SARS-CoV-2 and augment primary decontamination strategies such as ventilation.” Curtius, Granzin and Schrod (2021) reached similar conclusions based on measurements of aerosol concentration reductions in a classroom. Additionally, portable HEPA filters have been shown to significantly reduce concentrations of traffic-related aerosol concentrations in homes close to highways (Cox et al. 2018), improve clinical manifestations for patients with allergic rhinitis by reducing particulate matter and dust mite allergen concentrations in bedroom air (Luo et al. 2021), and reduce woodsmoke particles in wood-burning communities with measurable health benefits in relatively young and healthy subjects (Allen et al. 2011).

The cost of HEPA filter systems generally scales with their capacity, usually characterized by their clean air delivery rate (CADR) (Association of Home Appliance Manufacturers (AHAM) 2014). The CADR determines the number of equivalent air changes per hour (ACH) achievable in a room of a given size. For example, the typical floor size of a U.S. classroom is about 1,000 ft^2^ (93 m^2^) and with a volume of about 8,000 ft^3^ (227 m^3^). To achieve three ACH in a room this size, for example, requires a CADR of 400 ft^3^ min^−1^ (680 m^3^ h^−1^). AHAM recommends that the CADR of an air filter is about two-thirds of the room floor area, corresponding to a CADR of 666 ft^3^ min^−1^ for a 1,000 ft^2^ classroom. In the context of airborne infectious disease transmission, the risk of long-range transmission continually decreases as the CADR increases (Shen et al. 2021). Limitations to in-room filtration include noise, energy consumption, and initial and maintenance costs for replacement filters. An initial cost-survey of commercially available Energy Star rated in-room filters (U.S.EPA 2021a) designed for the residential market found costs ranging from $0.71 to $2.66 per CADR in units of ft^3^ min^−1^ (Pistochini 2021), making them inaccessible to many people and in many contexts.

A recently proposed, easy-to-construct, and low-cost alternative air filter constructed from MERV-13 filters and a box fan provides an opportunity for more people to access air filters in an affordable manner. This do-it-yourself (DIY) air filter, known as the “Corsi-Rosenthal Box” (hereafter, CR Box), is finding use in classrooms and other indoor environments across the U.S. through a grassroots movement driven by social media and the accessibility of the materials (Emanuel 2021). Although MERV-13 filters have a lower intrinsic filtration efficiency than HEPA filters, in-room air filtration using MERV-13 filters will still lead to a reduction in particle concentrations. While some work on airflow optimization in the CR Box has been done (Elfstrom 2021) and some initial characterization exists (Srikrishna 2021; Wieingartner, Rüggeberg and Wipf 2021), no systematic evaluation of the performance yet exists. Given the adoption of the CR Box in classrooms and other environments, such evaluation is critical.

Here, we characterize the Corsi-Rosenthal Box performance via measurement of size-dependent particle decay for particles >0.5 microns in a classroom and a home office with and without the CR Box operating. Our method allows for determination of CADR values above the 450 ft^3^ min^−1^ (765 m^3^ h^−1^) upper-limit of the standard method (Association of Home Appliance Manufacturers (AHAM) 2014). We compare the results for the CR Box to those measured for two commercial HEPA filters in terms of overall efficacy and cost.

## 2 Materials and Methods

Here, we provide an overview of the methods used, with full details in the Supplementary Material. Decay rates of salt particles (**Figure S1**) introduced to two rooms—a furnished but not occupied 5926 ft^3^ (168 m^3^) classroom and a 1277 ft^3^ (36.2 m^3^) furnished but not occupied home office—were measured with and without the air filters turned on. A box fan oriented at the wall operated at low speed throughout the measurements to maintain similar turbulence and mixing conditions between experiments. The measurements with this mixing fan on but the air filters turned off provides the baseline ventilation plus particle deposition rate, as these are the primary loss pathways for particles in a room. The measurements with the air filters on additionally include the influence of the filter. The effective air changes per hour (*ACH*) (actual air exchange + particle deposition to indoor surfaces + particle removal by an air cleaner) for each experiment were determined by fitting an exponential decay curve with a y-offset (*y*0) to the particle concentration (*N*_p_) during the decay period:

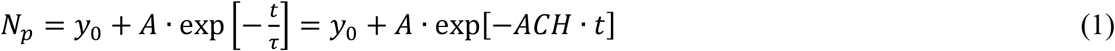

where *t* is the time in hours, *τ* is the decay lifetime in hours, and *A* is the amplitude.

When the air filters are turned on there is additional turbulence induced by the air filter fan that could alter the baseline deposition rate above that with the mixing fan alone. To assess the influence of this added turbulence we conducted experiments using two fans, the mixing fan and an additional box fan set in the location of the air filter. These experiments indicated that the additional turbulence from the air filter fan increased the baseline natural ventilation rate by 17 ± 11% in the home office but only 3% ± 3% in the classroom (**Figure S2**). The difference results from the classroom having active ventilation and a substantially higher baseline *ACH* compared to the home office (Table S1).

The *ACH* from filtration (*F*), ventilation (*V*), and deposition (*D*) add in series. Therefore, the *ACH* attributable to only the air filters (*ACH*_F_) is simply the difference between the value measured with the air filter on (*ACH*_F+V+D_) and the baseline *ACH* from room ventilation and particle deposition (*ACH*_V+D_):

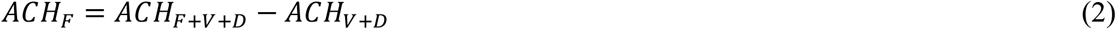

The *ACH*_V+D_ values used in Eqn. 2 are taken as the values measured with the air filter off and the mixing fan operating, but adjusted upwards by 17% or 3% to account for additional turbulence from the air filter fan. Eqn. 2 can be used to determine the weighted-average *ACH*_F_ across all particle sizes (by fitting to the particle number or mass concentration) or for specific size ranges. The corresponding CADR is:

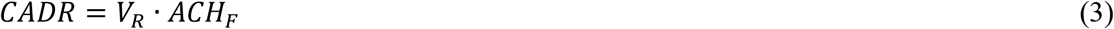

where *V*_R_ is the room volume. We use *ACH*_X*,N*p_ and *CADR*_*N*p_ when referring to the value determined from the particle number concentration and *ACH*_X,*M*p_ and *CADR*_*M*p_ when determined from the mass concentration, and where *X* corresponds to *F, V*, or *D* (or combinations thereof).

Particle concentrations and decay rates were measured at 5 s time resolution using an aerodynamic particle sizer (APS; TSI model 3321) and a low-cost sensor (LCS; Plantower PMS 5003). The APS characterizes particles into bins from 0.5-20 microns diameter according to their aerodynamic diameters (*D*_pa_) and thus allows for determination of size-specific *ACH* values. Size-specific values are only considered up to *D*_pa_ = 5.425 μm as above this value the decays are too noisy to allow for robust fitting, owing to the very low concentrations of particles above this size. The APS yields both number and mass concentrations, with the latter determined assuming a particle density of 2 g cm^−3^, consistent with the use of NaCl particles. Note that, unless otherwise stated, results are reported based on the APS measurements. The LCS converts light scattering observations to report size-dependent particle mass and particle number using an unknown algorithm with a nominal lower diameter limit of 0.3 microns. The reported number concentrations observed here exhibit linear decays (after natural log transformation), as expected, whereas the mass concentrations from the LCS exhibit distinctly non-linear decays. We therefore consider only the number concentration data from the LCS.

Three air filters were tested: the Corsi-Rosenthal Box and two commercial HEPA filters. The Corsi-Rosenthal Box was originally proposed by Richard Corsi on Twitter and with Jim Rosenthal making the first prototype (Rosenthal 2020). The CR Box used here is constructed using three 20” × 20” × 2” and two 16” × 20” × 2” MERV-13 filters (Air Handler, LEED/Green Pleated Air Filter, total cost $34.75) and a 20” box fan (Air King Model 4CH71G, $23.68). (See **Figure S3** and the Supplemental Material for a full description and discussion of cost). The CR Box here sits on legs that hold it about 4” (10 cm) off the ground and with the fan pointed upwards or sideways. In one variation, we tested the CR Box inverted such that the fan pointed at the floor, sitting about 4” (10 cm) off the floor. An inverted CR Box would potentially be more robust against potential foreign objects being dropped into the fan. One of the HEPA filters (HEPA #1) has a stated tobacco smoke CADR = 300 ft^3^ min^−1^ (510 m^3^ h^−1^) when operated at maximum speed while the other (HEPA #2) has a stated tobacco smoke CADR = 141 ft^3^ min^−1^ (240 m^3^ h^−1^) when operated at maximum speed.

The loudness of the air filters and of the box fan alone were measured using a decibel monitor (Extech Instruments HD600) that was situated 5 ft (1.52 m) from the center of the air filters and located perpendicular to the air exhaust. The power use by the air filters was measured using a power logger (Fluke 1735 Power Logger Analyst). Estimates of the fan airflow rate alone and as part of the CR box were estimated from air flow velocity measurements.

## 3 Results and Discussion

Example particle decays from the APS measurements for the natural room ventilation and with the various filters on are shown in **Figure 1**. The *ACH*_V+D_ was 3.5 ± 0.2 hr^−1^ (1σ, precision-based uncertainty) for the unoccupied classroom and the *ACH*_V+D_ was 1.3 ± 0.1 h^−1^ for the unoccupied home office. **Figure 2** shows the resulting clean air delivery rates for air cleaners that were tested in both the classroom and home office. Replicate CADR values for each air filter exhibited only small variations within each room and across the two rooms, although were generally larger for the measurements made in the classroom. The difference between the *ACH* values with and without the filter on was greater for the smaller home office (**Table S1**). Further, individual *ACH*_V+D_ values were determined for every air filter measurement in the home office but not the classroom (see Methods). Therefore, we take the *CADR* values determined from the home office as generally more reliable and, unless otherwise stated, use them in the discussion that follows.

**Figure 1.**
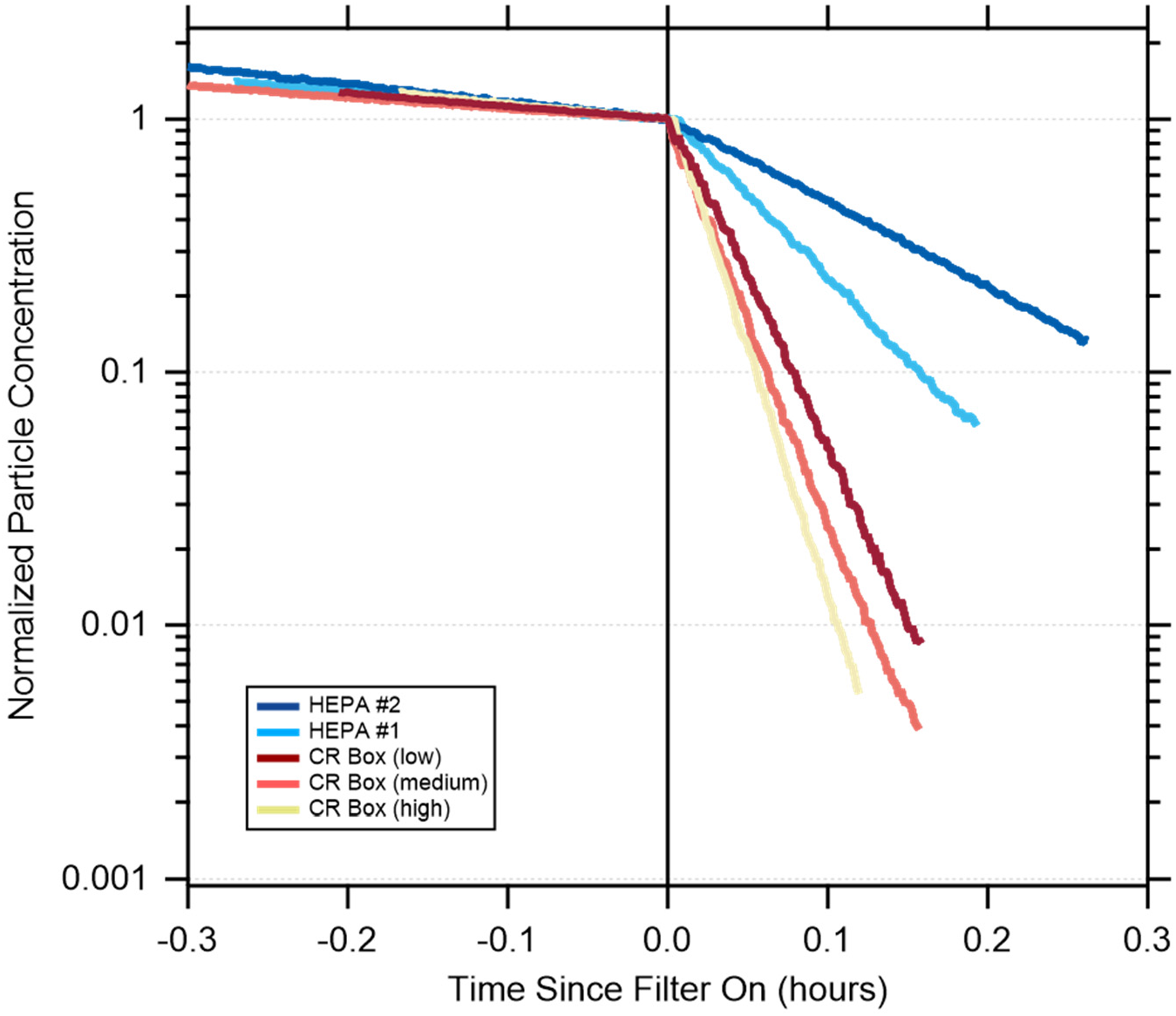
Example particle number decays measured in the home office for the natural ventilation + particle deposition (at *t* < 0) and with the air filters on (*t* > 0). Particle concentrations have been normalized to unity at *t* = 0.

**Figure 2.**
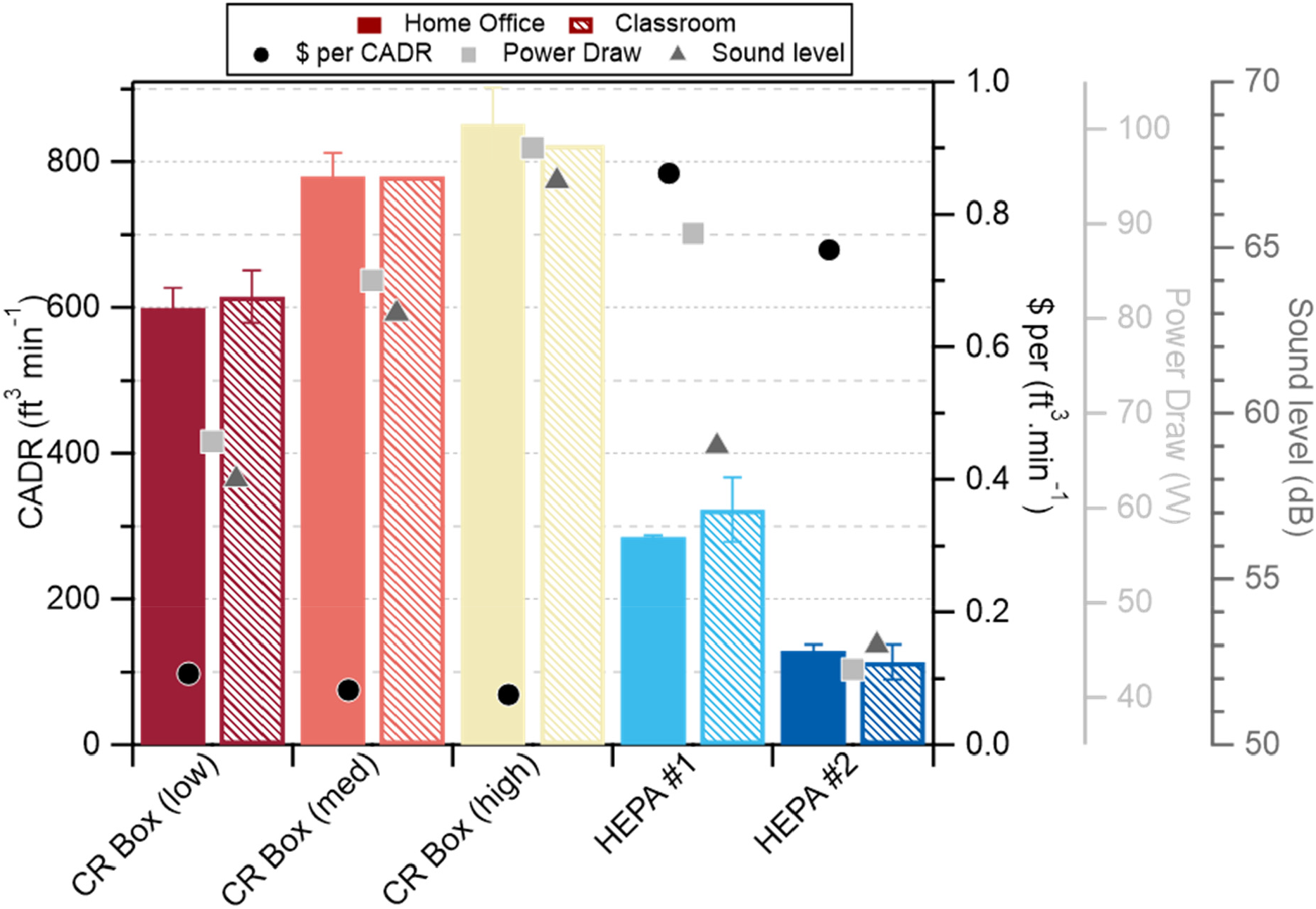
(left axis) The number-weighted clean air delivery rate for the various air filters (left axis, bars) as measured in the home office (left hash marks) and classroom (right hash marks). (right axis) The price normalized CADR (black circles), sound level (dark gray triangles), and power (light gray squares).

Generally, the *CADR*_Mp_ > *CADR*_Np_ with the exception of HEPA #2 (**Table S1**). For HEPA #1 the *CADR*_Np_ = 322 ± 44 ft^3^ min^−1^ (547 ± 75 m^3^ h^−1^) for the classroom and 285 ± 2 ft^3^ min^−1^ (484 ± 3.4 m^3^ h^−1^) for the home office, both in very good agreement with the manufacturer’s specification of 300 ft^3^ min^−1^ (510 m^3^ h^−1^). For HEPA #2 the *CADR*_Np_ = 113 ± 24 ft^3^ min^−1^ (192 ± 41 m^3^ h^−1^) for the classroom and *CADR*_Np_ = 129 ± 8 ft^3^ min^−1^ (219 ± 14 m^3^ h^−1^) for the home office, also in very good agreement with the manufacturer’s specification of 141 ft^3^ min^−1^ (240 m^3^ h^−1^). The good agreement between the measured *CADR*_Np_ and the manufacturer’s specifications provides a validation of the method.

The *CADR*_Np_ for the Corsi-Rosenthal Box increases reasonably linearly with fan speed (**Figure S4**), from 600 ± 27 ft^3^ min^−1^ (1020 ± 46 m^3^ h^−1^) at low speed to 852 ± 50 ft^3^ min^−1^ (1450 ± 85 m^3^ h^−1^) at high speed, as measured for the home office, and from 615 ± 36 ft^3^ min^−1^ (1045 ± 61 m^3^ h^−1^) to 823 ft^3^ min^−1^ (1400 m^3^ h^−1^) for the classroom. A linear fit with zero intercept to the *CADR*_Np_ for the home office versus the fan total airflow rate estimates for the box fan at the three speeds indicates an effective filter efficiency of 41-58%, with the range indicating uncertainty in the CR Box airflow rates (see Supplementary Material; **Figure S4**). The air velocity measurements with the filters added indicated a 12% reduction in flow with a pressure drop of 6.4 Pa. Accounting for this flow reduction increases the effective filter efficiency to 47-67%.

The size-dependent efficiency curves for the Air Handler MERV 13 filters indicates a minimum filtration efficiency (*η*_*f*_) of ∼55% for 0.35 micron diameter particles (*D*_p_), which increases to ∼85% for 0.75 micron diameter particles and to ∼90% for 1 micron dimeter particles (Air Handler via Grainger: 2022). Multiplying the observed particle size distribution by 1 - *η*_*f*_(*D*_*p*_) and comparing with the original particle size distribution indicates an expected size-averaged filtration efficiency of about 87% by number and 93% by mass, larger than observed. This difference may result from a much lower air velocity across the five parallel filters in this study (<148 ft min^−1^ = 1.07 m s^−1^) relative to those typically used for HVAC filter testing to determine MERV ratings (492 ft min^−1^ = 2.5 m s^−1^) (ASHRAE 2017). For MERV-13 filters, inertial impaction and interception are the dominant loss mechanisms for the size range of particles considered here (Flagan 1988). For these mechanisms, the single-fiber collection efficiency for fibers in a filter bed increases with the Stokes number, and therefore face velocity, and depends on the particle-to-fiber diameter ratio and fiber packing density. The Stokes number for a 1 micron diameter particle having a density of 1 g cm^−1^ encountering a 5 micron diameter fiber, fairly typical of modern filters (Kowalski and Bahnfelth 2002; Kowalski, Bahnfelth and Whittam 1999), at a face velocity of 492 ft min^−1^ (2.5 m s^−1^) equals 3.58, which is in the range over which the single-fiber filtration efficiency is particularly sensitive to changes in velocity (Flagan 1988). As such, a lower face velocity should mean lower removal efficiency due to the lessened effect of inertial impaction and interception. Alternatively, the reduced filtration efficiency measured in the experiment could also be attributed to leaks around the filter media, although the filter assembly was taped and visually inspected to seal any openings and thus we suspect that leaks play a minor role.

Size-dependent *ACH* and *CADR* values are determined by fitting decay curves to each particle size bin from the APS. The *ACH* for the natural room decay periods increase substantially with particle size (**Figure 3a**), likely due to higher particle deposition rates to indoor materials at larger aerodynamic diameters (Hussein and Kulmala 2008). The *ACH* for the air filters also increase with particle size, but to a lesser extent than the natural room decay (**Figure 3a**). Consequently, the *CADR* for the air filters, which derive from the difference between the filter on and natural room decay *ACH* values, exhibit a weaker dependence on particle size compared to the *ACH* (**Figure 3b**). The *CADR* for the CR Box vary only weakly with particle size for all speeds and are relatively constant from about 0.7 to 2.5 microns (**Figure 3b**), which is somewhat unexpected given the MERV-13 filter efficiency should increase sharply above 700 nm to about 1 micron, above which it should be constant and near unity. It is possible that the low face velocities on the filters relative to standard test conditions (ASHRAE 2017) led to atypical size dependence. Alternatively, additional turbulence from the filter exhaust air could have altered the particle deposition rates in the room from the baseline measurements leading to a flatter than expected size dependence, although the measurements with the added fan in place of the CR box suggest this had negligible influence. Notably, comparison of the size-specific *CADR* for particles with *D*_p,a_ > 1 μm to the specified fan speeds indicates an *η*_*f*_ much less than unity, even after accounting for the 12% reduction in flow owing to filter resistance. The reason for this apparent lower than expected *η*_*f*_ for the CR Box is unclear.

**Figure 3.**
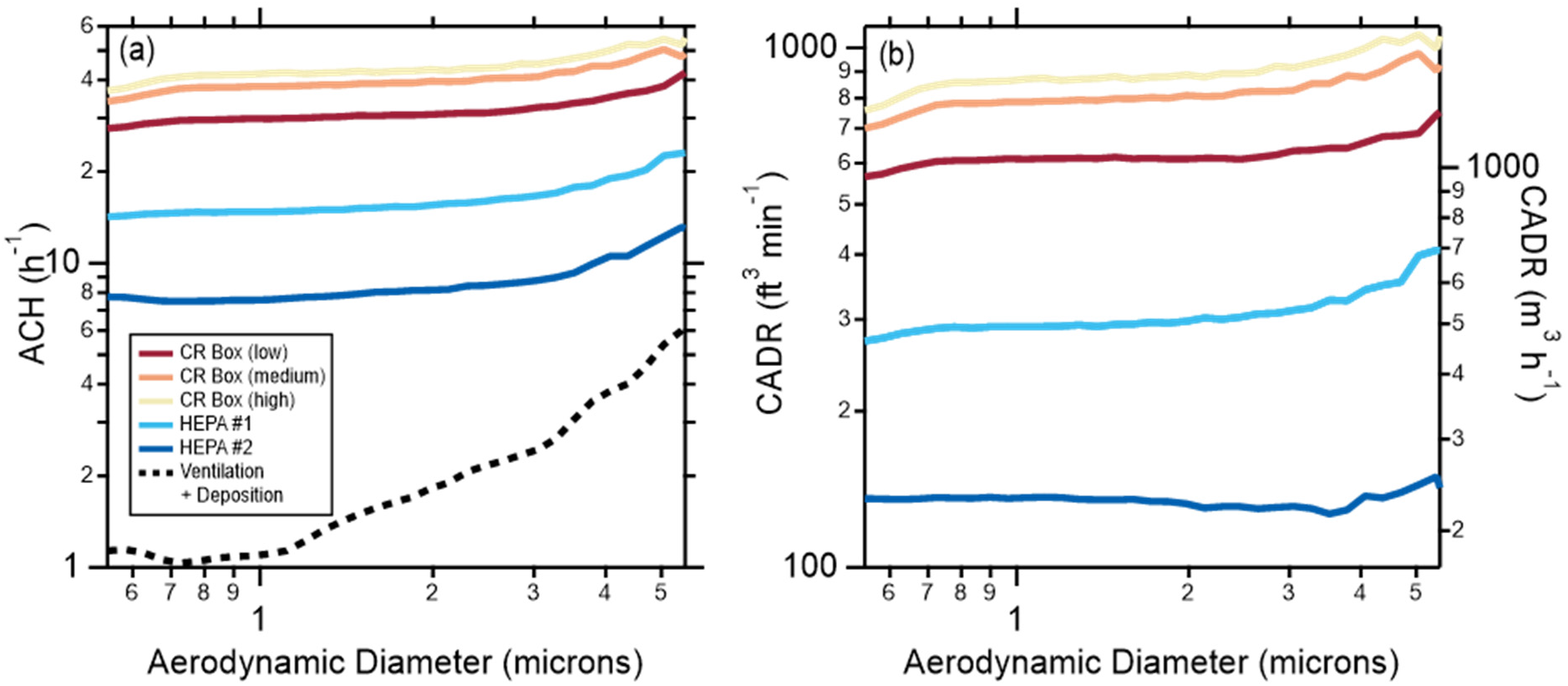
(a) Size-dependent air changes per hour with the various air filters operating (solid lines) and for the natural room ventilation + particle deposition alone (dashed line), as measured in the home office. (b) The corresponding clean air delivery rates for the various air filters. Results for the classroom are similar (not shown).

The CADR values for the Corsi-Rosenthal Box substantially exceed those of the particular commercial HEPA filters used here (Figure 2a). For further comparison, no U.S. Energy Star certified air cleaners have CADR values (for either tobacco smoke, dust, or pollen) matching the CADR value for the CR Box even on low speed (**Figure S5**). Consideration of the cost-per-unit-air-cleaned for the low-speed CR Box (<$0.072/(ft^3^ min^−1^)) and for the two HEPA filters (>$0.7/(ft^3^ min^−1^)) demonstrates that the DIY air filter is approximately one-tenth the initial cost of a commercially available HEPA filter per unit of air cleaned (**Figure 2**).

The CR Box loudness varied from 58 dB (low speed) to 67 dB (high speed) (**Figure 2**; **Table S1**). The low speed loudness is similar to that measured for HEPA #1 (59 dB) but higher than that for HEPA #2 (54 dB). For reference, a modern refrigerator has a noise rating of about 50 dB and a LEED certified vacuum must be <70 dB. To attain a CADR equivalent to the CR Box on low speed would require about two HEPA #1 units and 4 HEPA #2 units, which would yield 62 dB and 60 dB, respectively. The power draw for the CR Box varied from 67 W (low speed) to 98 W (high speed) and was 89 W for HEPA #1 and 43 W for HEPA #2, corresponding to 8.9 and 8.7 ft^3^ min^−1^.W^−1^ (15.1 and 14.8 m^3^ h^−1^.W^−1^) for the CR Box and 3.2 and 3.0 ft^3^ min^−1^.W^−1^ (5.4 and 5.1 m^3^h^−1^.W^−1^) for the HEPA filters. For comparison, the most efficient category of U.S. Energy Star certified portable air cleaners must have an efficiency equal or greater than 2.9 ft^3^ min^−1^.W^−1^ (4.9 m^3^ h^−1^.W^−1^, meaning the CR Box is three times more efficient than the Energy Star standard (U.S.EPA 2021b).

The *CADR* values for the inverted CR Box were all suppressed relative to the standard CR Box orientation (with the fan pointed upwards; **Table S2**). For example, in the inverted orientation the *CADR*_Np_ = 481 ft^3^ min^−1^ (817 m^3^ h^−1^) on the low setting, compared to ∼600 ft^3^ min^−1^ (1019 m^3^ h^−1^) in the standard orientation. This difference likely resulted from one or both of (i) short circuiting of the airflow wherein clean air exhausted by the fan is preferentially entrained into CR Box filters rather than being dispersed into the broader room, or (ii) increased shear forces that resuspended particles previously deposited on the floor and increase particle number concentrations in air. Therefore, we suggest that orienting a CR Box (or likely any air filter) such that the fan exhaust is towards the floor be avoided.

The CADR measurements made with the low-cost sensor yield generally similar results to those made with the APS (**Figure S6**), with the *CADR*_Np_ increasing with fan speed for the CR Box and with values for the HEPA filters similar to the manufacturer’s specification. However, the specific *CADR*_Np_ depended on which reported particle size regime was used for the fitting. The values determined for the *N*_p,>1.0_ bin generally agreed best with the *CADR*_Np_ from the APS, while the *N*_p,>0.3_ and *N*_p,>0.5_ values were generally smaller and the *N*_p,>2.5_ and *N*_p,>5.0_ values were larger. Without further knowledge of the algorithm behind the low-cost sensor data processing we cannot establish the origin of this apparent size dependence or why the *N*_p,>1.0_ bin yields the most similar values. Nonetheless, our results suggest that the use of low-cost sensors can yield a reasonable measure of the relative *CADR* values between air filters and a reasonable estimate of the absolute *CADR* values, and thus a means by which those without access to expensive instrumentation can determine the efficacy of DIY air filters.

## 4 Summary

We have measured the filtration efficiency for particles >0.5 microns of a DIY, open-source air filtration system, the Corsi-Rosenthal Box, comprised of a box fan and MERV-13 filters. At the lowest speed the clean air delivery rate is >600 ft^3^ min^−1^ (1019 m^3^ h^−1^), demonstrating exceptional performance relative to most commercially available air filters. The CADR increases with fan speed, with the highest value about 850 ft^3^ min^−1^ (1444 m^3^ h^−1^). However, the filter noise level also increases with fan speed, from 58 dB at low speed to 67 dB at high speed. The CR Box is cost efficient, with a cost-normalized CADR of <$0.072/(ft^3^ min^−1^). Future efforts to improve and characterize the CR Box might focus on decreasing the CR Box noise level without compromising filtration performance or characterizing different CR Box designs that use different fans and filters or different numbers of filters.

## Data Availability

All data produced in the present study are available upon reasonable request to the authors

## 5 Acknowledgements

The authors greatly appreciate the contributions from students in ECI/ATM 149 at UC Davis, who performed some preliminary experiments as part of a lab activity. Jim Rosenthal is thanked for constructing and promoting the first Corsi-Rosenthal box.

## Author contributions

RDP, MNK, TP, and CDC made measurements; RDP and CDC analyzed data; all authors interpreted results; RC designed the DIY air filter; CDC and RDP led the writing with contributions from all authors.

## Data and materials availability

All data needed to evaluate the conclusions in the paper are present in the paper and/or the supplementary materials.

## References

Air Handler via Grainger: (2022), Green pleat merv 13, https://www.grainger.com/ec/pdf/Air-Handler-Green-Pleat-Merv-13-Air-Filters-Guide.pdf, Accessed: 2 January 2022.

Alavy, M. and J. A. Siegel. 2020. In-situ effectiveness of residential hvac filters. Indoor Air 30:156–166. doi: 10.1111/ina.12617.

Allen, R. W., C. Carlsten, B. Karlen, S. Leckie, S. van Eeden, S. Vedal, I. Wong, M. Brauer. 2011. An air filter intervention study of endothelial function among healthy adults in a woodsmoke-impacted community. American Journal of Respiratory and Critical Care Medicine 183:1222–1230. doi: 10.1164/rccm.201010-1572OC.

ASHRAE. 2017. Standard 52.2: Method of testing general ventilation air-cleaning devices for removal efficiency by particle size. Atlanta, GA.

Association of Home Appliance Manufacturers (AHAM). 2014. Ansi/aham ac-1: Method for measuring the performance of portable household electric room air cleaners. Washington, DC.

Cox, J., K. Isiugo, P. Ryan, S. A. Grinshpun, M. Yermakov, C. Desmond, R. Jandarov, S. Vesper, J. Ross, S. Chillrud, K. Dannemiller, T. Reponen. 2018. Effectiveness of a portable air cleaner in removing aerosol particles in homes close to highways. Indoor Air 28:818–827. doi: 10.1111/ina.12502.

Curtius, J., M. Granzin, J. Schrod. 2021. Testing mobile air purifiers in a school classroom: Reducing the airborne transmission risk for sars-cov-2. Aerosol Science and Technology 55:586–599. doi: 10.1080/02786826.2021.1877257.

Elfstrom, D. (2021), by @DavidElfstrom, Title, Posted on: 15 August 2021, https://twitter.com/DavidElfstrom/status/1427112878616817669.

Emanuel, G. (2021), Does your kid’s classroom need an air purifier? Here’s how you can make one yourself, NPR, Published on: 26 August 2021, https://www.npr.org/sections/back-to-school-live-updates/2021/08/26/1031018250/does-your-kids-classroom-need-an-air-purifier-heres-how-you-can-make-one-yoursel.

Flagan, R. C. 1988. Fundamentals of air pollution engineering. Englewood Cliffs, N.J.: Prentice-Hall.

Hussein, T. and M. Kulmala. 2008. Indoor aerosol modeling: Basic principles and practical applications. Water, Air, & Soil Pollution: Focus 8:23–34. doi: 10.1007/s11267-007-9134-x.

Kelly, F. J. and J. C. Fussell. 2019. Improving indoor air quality, health and performance within environments where people live, travel, learn and work. Atmospheric Environment 200:90–109. doi: 10.1016/j.atmosenv.2018.11.058.

Kowalski, W. J. and W. P. Bahnfelth (2002), Merv filter models for aerobiological applications, https://www.nafahq.org/merv-filter-models/, Accessed: 2 January 2022.

Kowalski, W. J., W. P. Bahnfelth, T. S. Whittam. 1999. Filtration of airborne microorganisms: Modeling and prediction. ASHRAE Transactions: Research 105:4-17. doi.

Liu, D. T., K. M. Phillips, M. M. Speth, G. Besser, C. A. Mueller, A. R. Sedaghat. 2021. Portable hepa purifiers to eliminate airborne sars-cov-2: A systematic review. Otolaryngol. Head Neck Surg.:8. doi: 10.1177/01945998211022636.

Luo, J. Y., L. L. Ou, J. Ma, X. Y. Lin, L. M. Fan, H. C. Liu, B. Q. Sun. 2021. Efficacy of air purifier therapy for patients with allergic asthma. Allergol. Immunopath. 49:16–24. doi: 10.15586/aei.v49i5.146.

McNamara, M. L., J. Thornburg, E. O. Semmens, T. J. Ward, C. W. Noonan. 2017. Reducing indoor air pollutants with air filtration units in wood stove homes. Science of the Total Environment 592:488–494. doi: 10.1016/j.scitotenv.2017.03.111.

MillerLeiden, S., C. Lobascio, W. W. Nazaroff, J. M. Macher. 1996. Effectiveness of in-room air filtration and dilution ventilation for tuberculosis infection control. Journal of the Air & Waste Management Association 46:869–882. doi: 10.1080/10473289.1996.10467523.

Pistochini, T. (2021), Considerations for use and selection of portable air cleaners for classrooms, Rep., 3 pp, Western Cooling Efficiency Center, University of California, Davis, https://ucdavis.app.box.com/s/81yd5wsylxsc8oi2vtgf569vorph3tfk.

Rosenthal, J. (2020), A variation on the “box fan with merv 13 filter” air cleaner, https://www.texairfilters.com/a-variation-on-the-box-fan-with-merv-13-filter-air-cleaner/, Accessed: 2 January 2022.

Shaughnessy, R. J. and R. G. Sextro. 2006. What is an effective portable air cleaning device? A review. Journal of Occupational and Environmental Hygiene 3:169–181. doi: 10.1080/15459620600580129.

Shen, J. L., M. Kong, B. Dong, M. J. Birnkrant, J. S. Zhang. 2021. Airborne transmission of sars-cov-2 in indoor environments: A comprehensive review. Sci. Technol. Built Environ. 27:1331–1367. doi: 10.1080/23744731.2021.1977693.

Srikrishna, D. 2021. Price-performance comparison of hepa air purifiers and lower-cost merv 13/14 filters with box fans for filtering out sars-cov-2 and other particulate aerosols in indoor community settings. preprint on medRxiv. doi: 10.1101/2021.12.04.21267300.

Taber, C. and M. Ivanovich. 2018. New federal regulations for ceiling fans. ASHRAE Journal 60:42-46. doi.

U.S.EPA (2021a), Energy star certified air purifiers (cleaners), https://www.energystar.gov/productfinder/product/certified-room-air-cleaners/results, Accessed: 29 December 2021.

U.S.EPA (2021b), Energy star product specifications for room air cleaners: Eligibility criteria version 2.0, https://www.energystar.gov/sites/default/files/ENERGY%20STAR%20Version%202.0%20Final%20Room%20Air%20Cleaners%20Program%20Requirements_0.pdf, Accessed: 29 December 2021.

Wieingartner, E., T. Rüggeberg, M. Wipf (2021), Measurement of the filtration performance of diy air cleaners (cadr values) for aerosol particles with diameters smaller than 1 micrometer., Rep., 13 pp, University of Applied Sciences Northwestern Switzerland, https://makehumantechnology.org/wp-content/uploads/2021/10/Messbericht_DIY_Air_Cleaner_2021-10-8-EN.pdf.

