## Supplemental Materials for "Characterizing the performance of a DIY air filter"

The supplemental materials contains:

- A fuller description of the materials and methods
- Tables S1-S2
- Figures S1-S6

### Materials and Methods

#### Particle Generation and Measurement

Our overall approach to determining air changes per hour, either with or without added air filtration, generally follows from the measurement of decay rates of particles introduced into rooms at concentrations well above background. Particles were generated using portable mesh nebulizers (Wellue®) filled with an aqueous solution of table salt (100 g $L^{-1}$). The nebulizers were operated on their maximum setting (0.9 ml $min^{-1}$) and up to two were used per room. A representative particle size distribution is shown in Figure S1. A box fan was turned on its low setting and positioned about 0.5 m from one wall of the room, pointing at the wall, to induce mixing in the room, ideally leading to a reasonably well-mixed condition.

Although a similar approach was taken for measurements made in the classroom and the home office, the experimental details differed slightly. For the classroom, experiments began with measurement of the background particle concentration with all doors to the room closed. Following the background measurement the nebulizer was turned on with the air filters turned off. The nebulizer automatically shut off after ~10 minutes at which point the air filters were turned on. Particle measurements continued for an additional ~30 minutes during which the particle decay was measured. The first experiment was conducted with the CR Box turned off to determine the baseline effective room air exchange rate owing to the ventilation or natural infiltration or particle deposition to surfaces. Subsequent experiments had the CR Box turned on to either the low, medium, or high setting or the HEPA filters turned on to their highest setting. For the classroom, two replicate measurements were made for each of the HEPA filters and for the CR Box on low speed, but only one measurement each was made for the CR box at medium and high speed. For the home office the protocol differed slightly. Specifically, following the background particle measurement and subsequent particle generation the decay from natural ventilation/infiltration plus deposition was measured for ~20 minutes. At this point the air filter of interest was turned on and the decay with the air filter on was measured for ~15 minutes. This allowed for determination of a unique baseline air changes per hour for every filter measurement. For the CR Box three replicate measurements at each speed were made, while only two replicates were made for the commercial HEPA filters in the home office.

The air exchange rate was determined by fitting an exponential decay curve with a y-offset ($y_0$) to the particle concentration ($N_p$) period starting approximately one minute after the nebulizer stopped, where:

$$N_p = y_0 + A \cdot \exp\left[-\frac{t}{\tau}\right] = y_0 + A \cdot \exp[-ACH \cdot t] \qquad (1)$$

where $t$ is the time in hours, $\tau$ is the decay lifetime, and $A$ is the amplitude. The $y_0$ is determined by the particle background concentration and the $A$ by the particle source rate relative to the overall air exchange rate. The background particle concentrations were sufficiently small that we could assume $y_0 = 0$ with no meaningful influence on the results. The $ACH$ attributable to only the CR Box ($ACH_F$) is simply the difference between the value measured with the CR Box on and the baseline $ACH$ from room ventilation and particle deposition (that is, with the filter off, $ACH_{V+D}$), as these add in series.

$$ACH_F = ACH_{F+V+D} - ACH_{V+D} \qquad (2)$$

The robustness of the fits from Eqn. 1 were verified via linear fitting to the natural log transformed and background-subtracted particle concentration data. Eqn. 1 can be used to determine the weighted-average $ACH_F$ across all particle sizes (by fitting to the particle number or mass concentration) or for specific size ranges. The corresponding CADR is:

$$CADR = V_R \cdot ACH_F \tag{3}$$

where $V_R$ is the room volume, and with appropriate unit conversion. We use $ACH_{X,N_p}$ and $CADR_{N_p}$ when referring to the value determined from the particle number concentration and $ACH_{X,M_p}$ and $CADR_{M_p}$ when determined from the mass concentration, and where $X$ corresponds to $V+D$ (natural room ventilation and deposition only), $F$ (filter only), or $F+V+D$ (filter + natural room ventilation + deposition).

The influence of additional turbulence induced by the fan in the CR Box on particle deposition to surfaces was also assessed for the classroom and the home office. In both, a single fan, oriented towards a wall, was first turned on similar to the filtration experiments above. The nebulizer was then started and particles were produced for about 10 minutes and the concentration of particles in the room increased. Once the nebulizer stopped the particle concentration in the room was allowed to decay for 10-20 minutes with just the single fan operating. Then, a second fan with no filters attached was started and the particle decay was measured for an additional 20-30 minutes. This second fan was placed in the same position and with the same orientation as the fan in the CR Box. Example decays are shown in Figure S2.

Particle concentrations and decay rates were measured using two independent methods. An aerodynamic particle sizer (APS; TSI model 3321) characterized particles having aerodynamic diameters from 0.5-20 microns with 5-second time resolution. The APS characterizes particles into bins according to their aerodynamic diameters ($D_{pa}$) and thus allows for determination of size-specific $ACH$ values. Size-specific values are only considered up to $D_{pa}$ = 5.425 μm as above this value the decays are too noisy to allow for robust fitting. The APS is a well-established instrument for the characterization of particle concentrations and size distributions. As such, we use the measurements made with the APS for the main analysis in the main text.

A low-cost Plantower sensor (PMS 5003) characterized particles having optical diameters above about 0.3 microns with 5-second time resolution. The Plantower sensor converts and reports observations of scattering to size-dependent particle mass and particle number using an algorithm that is unknown. Also unknown is the relationship between particle number and mass concentration. The reported number concentrations observed here exhibit linear decays (after natural log transformation) whereas the mass concentrations exhibit distinctly non-linear decays. The reason for this is unclear, as one would expect that the number concentration and mass concentrations are related through a simple linear transformation for this type of instrument. Regardless of reason, since the number concentration measurements exhibit a linear decay, similar to the APS, we consider only the number concentration data from the Plantower sensor.

**Air Filters**

Three air filters were tested: the Corsi-Rosenthal Box and two commercial HEPA filters.

***The Corsi-Rosenthal Box***

The Corsi-Rosenthal Box was originally proposed by Rich Corsi on Twitter and with Jim Rosenthal making the first prototype (Rosenthal 2020). The CR Box used here is constructed using three 20" x 20" x 2" and two 16" x 20" x 2" MERV-13 filters (Air Handler, LEED/Green Pleated Air Filter, total cost \$34.75) and a 20" box fan (Air King Model 4CH71G (9723), \$23.68). The assembled Corsi-Rosenthal Box is shown in Figure S3. We note that the cost of the filters here was about half that from many vendors, possible owing to purchasing agreements between UC Davis and specific vendors. In a non-comprehensive internet search conducted on 21 November 2021 we found that the average price for a MERV-13 20" x 20" x 2" filter averaged \$13.19 ± \$2.22 and for a 16" x 20" x 2" filter averaged \$15.39 ± \$3.96, corresponding to a total cost of \$70.36. Similarly, the particular Air King fan used here retails for about twice our purchase price when not on sale. Two of the 16" x 20" and two of the 20" x 20" filters are used to construct the side walls that sit on the 20" x 20" filter. The box fan is attached to the top and the seams are sealed with duct tape (\$6). The box fan is oriented such that the fan blows out of the constructed filter box. This creates a slight negative pressure that may help to seal the box and limit leaks, although any persistent leaks from e.g., holes in the filters or the tape would be independent of the flow direction. The box fan includes a ~circular "shroud" that covers the box fan corners and prevents backflow of unfiltered air into the fan. (The use of a shroud was proposed for square box fans by David Elfstrom on Twitter (Elfstrom 2021).) Here the diameter of the open shroud is 17". The CR Box sits on legs that hold it about 4" (10 cm) off the ground and with the fan pointed upwards or sideways. In one variation, we tested the CR Box inverted such that the fan pointed at the floor, sitting about 4" (10 cm) off the floor. An inverted CR Box would potentially be more robust against potential foreign objects being dropped into the fan.

*Commercial HEPA filters*

Two commercial HEPA filters were tested. One (HEPA #1) has a stated tobacco smoke CADR = 300 $ft^3$ $min^{-1}$ (508 $m^3$ $h^{-1}$) when operated at maximum speed and includes two prefilters to capture larger particles and reduce volatile organic compounds, and a HEPA filter. It retails for about $250. The other (HEPA #2) has a stated tobacco smoke CADR = 141 $ft^3$ $min^{-1}$ (240 $m^3$ $h^{-1}$) when operated at maximum speed, includes an activated carbon prefilter, and a noise level of 50 dB as specified by the manufacturer. It retails for about $100.

**Measurement Environment**

Measurements were made initially in three environments: (i) a 5926 $ft^3$ (167.8 $m^3$) classroom in Ghausi Hall at UC Davis; (ii) a 2890 $ft^3$ (81.8 $m^3$) office/meeting space in Ghausi Hall; and (iii) a 1277 $ft^3$ (36.2 $m^3$) home office in a residential building dating to 1923. For the home office the HVAC system was kept off throughout the measurements and thus the natural decay depended only on infiltration/exfiltration rates and particle deposition. Ultimately, only two of these environments were considered (the classroom and the home office) because the ventilation rates were sufficiently constant. In the office/meeting space the ventilation rate reduced when the occupancy sensors detected no movement for 15 minutes and shut off airflow after an additional 15 minutes of no movement. The ventilation rate in this room was too variable to allow for robust determination of the filter-specific *ACH* values. The smaller size of the home office compared to the classroom led to a greater difference in the *ACH* values measured with an air filter on versus with it off.

**Fan Speed & Measurement**

The fan has a manufacturer specified air flowrate of 1463, 1900, and 2163 $ft^3$ $min^{-1}$ (2486, 3228, 3675 $m^3$ $h^{-1}$) for low, medium, and high settings, respectively, tested under AMCA 230-99, which tends to overestimate fan speeds by 30% (Taber and Ivanovich 2018), although it is questionable how well this applies to box fans as the AMCA 230-99 method was developed for ceiling fans. Regardless, a 30% reduction corresponds to reduced air flowrates of 1024, 1330, and 1514 $ft^3$ $min^{-1}$ (1740, 2260, 2572 $m^3$ $h^{-1}$). The face velocities on the five filters (area ~ 10.25 $ft^2$ or 0.95 $m^2$) are 143, 185, and 211 ft $min^{-1}$ (0.75, 0.94, 1.07 m $s^{-1}$) using the manufacturer's values and 100, 130, and 148 ft $min^{-1}$ (0.51, 0.66, 0.75 m $s^{-1}$) using the reduced values. However, taping the corners of the box fans can also lead to an increase in the air flow rate through the filters as it reduces the potential for back flow.

To assess these estimates, we measured the air velocity in feet per minute using a Veloci Calc Model 9555-P. Measurements were made at six radial positions approximately equidistant from each other, starting at the center of the fan, moving to the outer edge. These six positions were measured at the mid-point of each edge of the fan. Measurements were first taken with the fan as purchased, with no modifications, at both high and low speeds. Then, the corners of the fan were taped, and a measurement at low speed was taken with no filters and with 5 filters in the Corsi-Rosenthal Box configuration. The face velocity was integrated over the 24 measurements and their distance from the center. Average face velocities for the unaltered fan at high speed were 880 ft $min^{-1}$ (268 m $min^{-1}$), 650 ft $min^{-1}$ (198 m $min^{-1}$) for the unaltered fan on low speed, 668 ft $min^{-1}$ (204 m $min^{-1}$) for the fan on low speed with taped edges, and 578 ft $min^{-1}$ (176 m $min^{-1}$) for the Corsi-Rosenthal on low speed. Multiplying by the fan area but without accounting for the area taken up by the fan protective grate, these equate to air flowrates of 1800 $ft^3$ $min^{-1}$ (3058 $m^3$ $h^{-1}$) for the high velocity fan with no modifications, 1331 $ft^3$ $min^{-1}$ (2261 $m^3$ $h^{-1}$) for the low setting with no modifications, 1361 $ft^3$ $min^{-1}$ (2312 $m^3$ $h^{-1}$) for the low setting with the fan edges taped, and 1171 $ft^3$ $min^{-1}$ (1990 $m^3$ $h^{-1}$) for the low speed in CR configuration. Accounting for the protective grate area would increase these slightly. These values for the unadulterated fan are in between the reported and reduced manufacturers' specified values, but within the likely uncertainties. The fan flow rate in the CR Box configuration with the fan edges taped is reduced by only 12% from the unaltered fan.

**Loudness & Current Measurement**

The loudness of the air filters was measured using a decibel monitor that was situated 5 ft (= 1.52 m) from the center of the air filters and located perpendicular to the air exhaust. The background room noise level was 40 dB. Measurements were also made for the box fan separate from the filters. The power draw by the air filters were measured using a Fluke power meter. Because dB is a logarithmic scale, noise levels ($L$) must be added after log transformation as:

$$L = 10 \cdot \log\left(\sum_{i=1}^{n} 10^{\frac{L_i}{10}}\right)$$

### Supplementary Tables

**Table S1:** Measured air changes per hour and clean air delivery rates.

| Air Filter | $ACH_{F+V+D}$ # ($h^{-1}$) | $CADR_{Np}$ * ($ft^3$ $min^{-1}$) | $ACH_{F+V+D}$ # ($h^{-1}$) | $CADR_{Np}$ * ($ft^3$ $min^{-1}$) | $CADR_{Mp}$ * ($ft^3$ $min^{-1}$) | Noise level (dB) | Power Draw (W) | $ per CADR |
|---|---|---|---|---|---|---|---|---|
| | ***Classroom*** | | | ***Home office*** | | | | |
| None | 3.5 | -- | 1.3 ± 0.14 | -- | | 40 | | |
| CR Box (low) | 9.8±0.4 | 614±36 | 29.6 ± 1.5 | 599 ± 27 | 614 ± 26 | 58 ± 2 | 67 | 0.11 |
| CR Box (med) | 11.4 | 780^ | 38.0 ± 0.1 | 780 ± 32 | 824 ± 32 | 63 ± 2 | 84 | 0.08 |
| CR Box (high) | 11.9 | 823^ | 41.5 ± 1.7 | 852 ± 50 | 903 ± 49 | 67 ± 1 | 98 | 0.08 |
| HEPA #1 | 6.8±0.4 | 323±44 | 15.4 ± 0.5 | 285 ± 2 | 300 ± 2 | 59 ± 1 | 89 | 0.86 |
| HEPA #2 | 4.7±0.3 | 114±24 | 7.9 ± 0.2 | 129 ± 8 | 118 ± 3 | 54 ± 1 | 43 | 0.74 |

#Based on number concentration measurement; not adjusted for additional turbulence

*Calculated from individual pairs of $ACH_{V+D}$ and adjusted $ACH_{F+V+D}$ and so may not match with the CADR determined from the average $ACH_{F+V+D}$

^Only one measurement was made

**Table S2:** Measured clean air delivery rates ($ft^3$ $min^{-1}$) for the Corsi-Rosenthal Box in the inverted orientation

| Air Filter | $CADR_{Np}$ ($ft^3$ $min^{-1}$) | $CADR_{Mp}$ ($ft^3$ $min^{-1}$) |
|---|---|---|
| CR Box (low) | 481 | 489 |
| CR Box (med) | 728 | 763 |
| CR Box (high) | 809 | 854 |

### Supplementary Figures

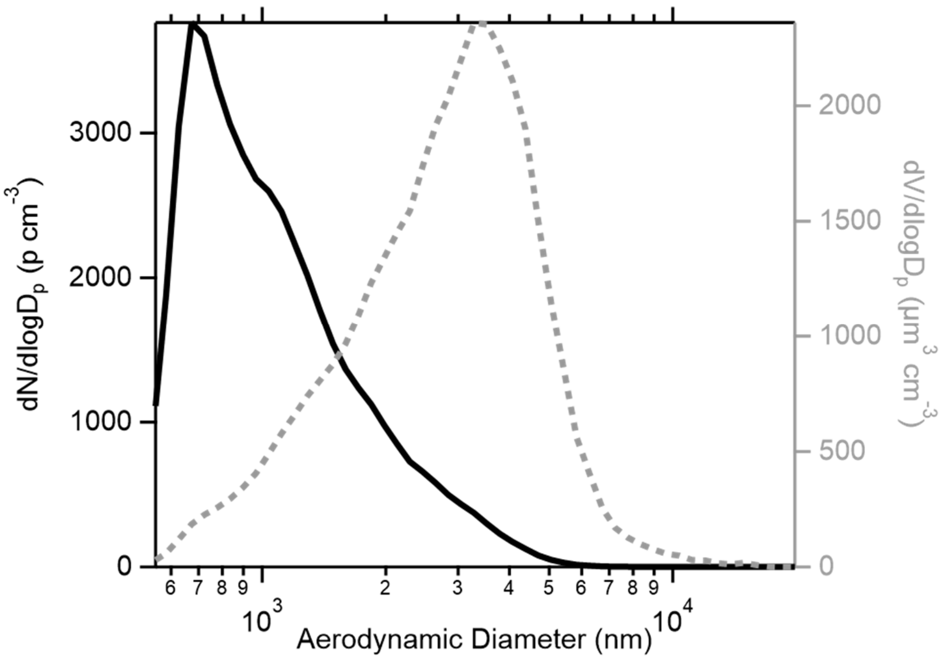


**Figure S1.** Number weighted (black solid, left axis) and volume weighted (gray dashed, right axis) particle size distributions for the particles produced from the nebulizer.

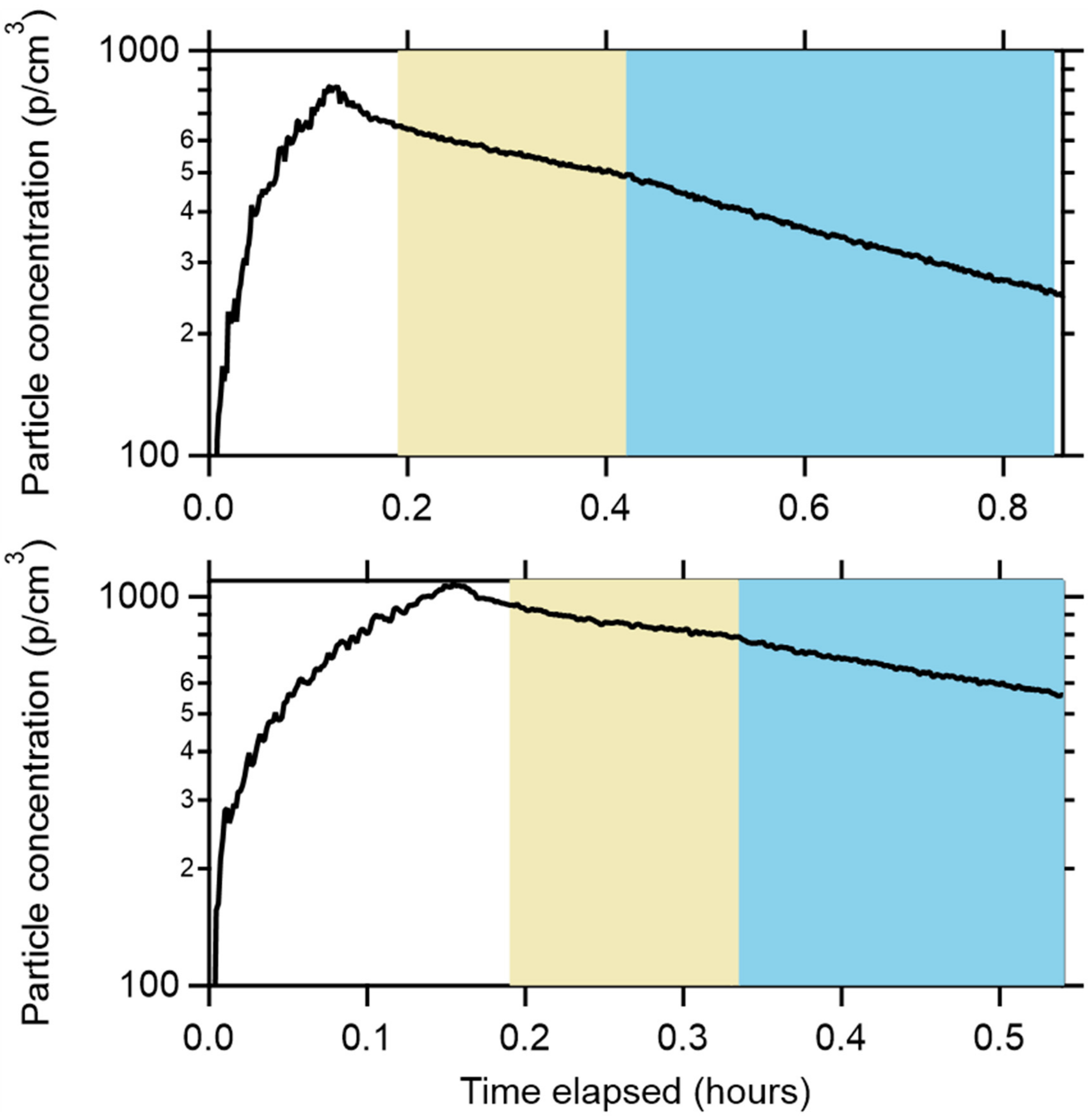


**Figure S2.** Particle decay with one fan (yellow) and with two fans (blue) for the home office (top) and classroom (bottom).

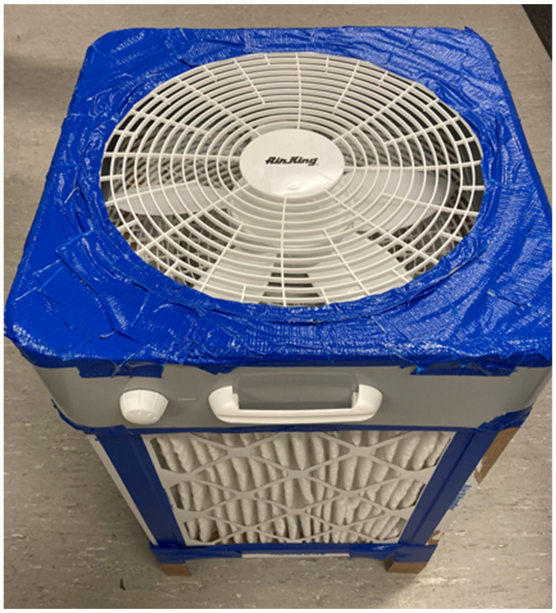


**Figure S3.** A photo of the assembled Corsi-Rosenthal Box.

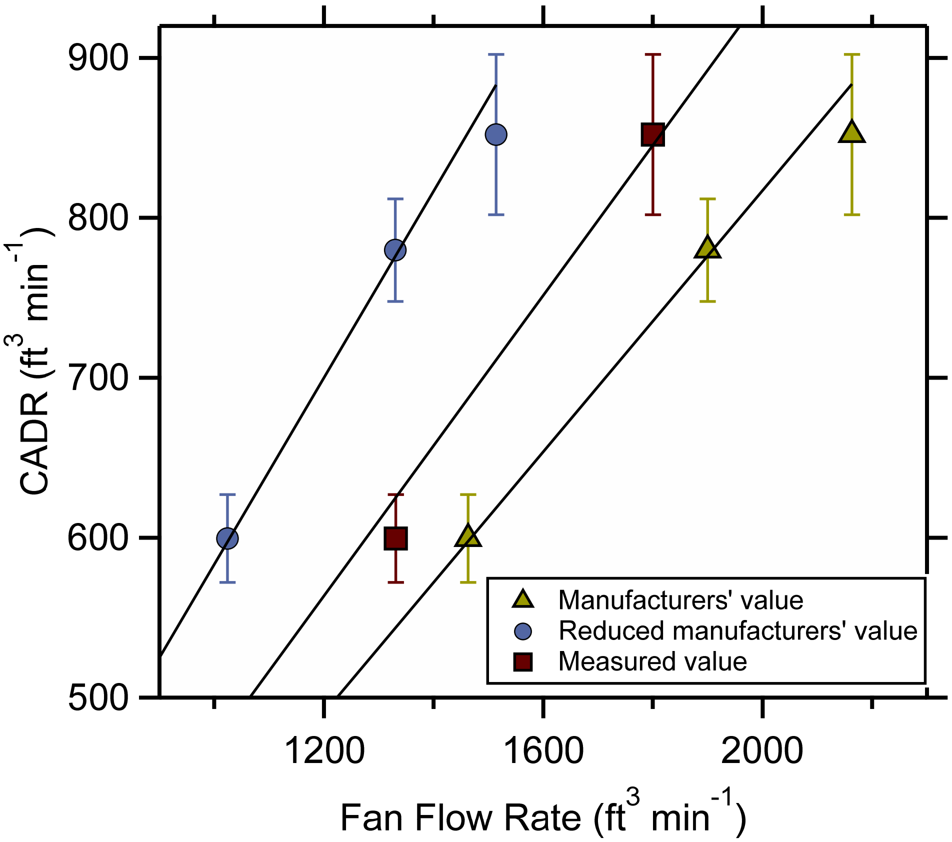


**Figure S4.** Relationship between CADR for the Corsi-Rosenthal Box and the manufacturer reported (yellow triangles), the reduced manufacturer reported (blue circles), and air-velocity-estimated (red squares) air flow rates for the original box fan. The lines are linear fits forced through zero (slopes = 0.41, 0.58, and 0.47, respectively).

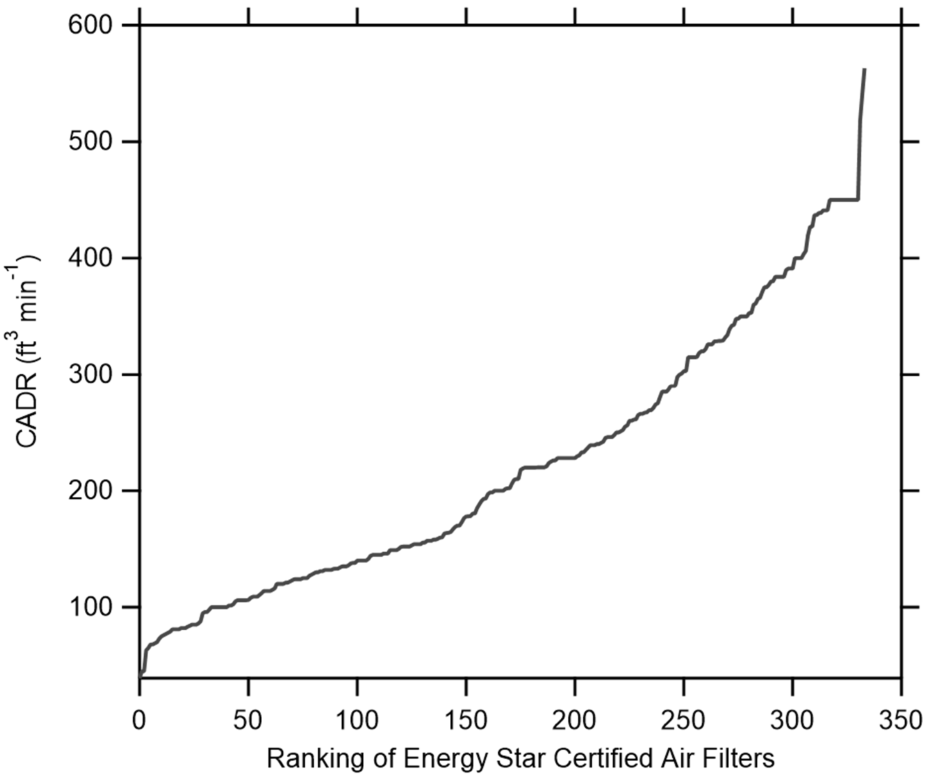


**Figure S5.** The distribution of maximum CADR values for commercially available air filters in the Energy Star database. These can be compared with the Corsi-Rosenthal Box.

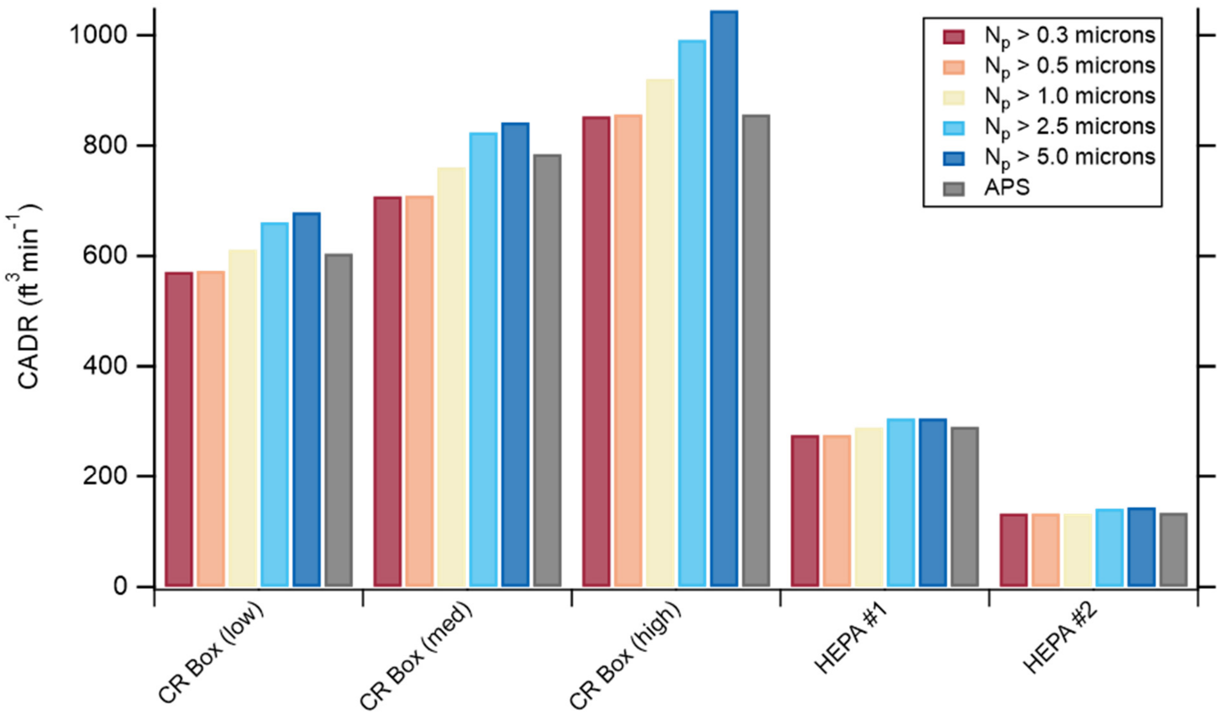


**Figure S6.** Comparison of the CADR values determined using the different apparent size bin number concentrations from the low-cost sensor (colors) compared to the $CADR_{Np}$ determined from the APS. Measurements are from the home office.
